# Fast-Imagery Reversal Script for Trauma-release (FIRST^®^): proof of concept in two populations with employment-related PTSD

**DOI:** 10.64898/2026.05.11.26352892

**Authors:** J Sturt, A Grealish, V Tzouvara, R Rogers, L De Rijk, C Armour, D Cameron, B Croak, Y Cui, F Fiorentino, R Harris, E Herrall, O Idowu, J Kreft, A Murray, V Pile, E Rowland, J Shephard, E Spikol, S Stevelink, H Strang, H Winter, A Wright-Hughes, N. Greenberg

**Affiliations:** King’s College London; University of Limerick; Queen’s University Belfast; Inspire Wellbeing; University of Leeds; South London and Maudsley NHS Foundation Trust; JSK Psychological Solutions

**Keywords:** Post traumatic stress disorder (PTSD), Veterans, Health and Social care workers, Therapy, Trial, Complex PTSD, Neurolinguistic programming (NLP)

## Abstract

**Background:** Post-Traumatic Stress Disorder (PTSD) is a mental health condition affecting people who experience traumatic events. Trauma-exposed occupational groups report higher rates of PTSD than the general population. Current treatments, and access, often take months and may cause distress when people are required to talk about the trauma.

**Objective:** To determine the proof of concept of FIRST, a brief, non-trauma focussed therapy, in two separate populations with employment-associated PTSD.

**Method:** Two independent, single-arm, experimental therapy pilot trials were conducted. Trial one recruited 20 military veterans who received FIRST therapy via trained third-sector therapists. Trial two recruited 16 health and social care workers with FIRST therapy delivered by healthcare provider therapists. All participants were adults with PTSD (confirmed via CAPS-5 in trial one, and symptom score of ≥33 on the PCL5 in trial two). Primary outcomes were recruitment feasibility, retention, data quality and reduction in PTSD symptoms. Secondary outcomes were anxiety and depression symptoms, daily life functioning and perceived health status. Veterans were followed up at 12 weeks post-enrolment and healthcare workers at 8 weeks.

**Results:** The veteran trial progression criteria to main trial were met. Seventy-nine people screened eligible, 43 attended a CAPS-5 assessment; 20 had confirmed PTSD and were enrolled. Seventeen completed therapy and 12-week outcome measures. Mean PCL-5 scores decreased from 48.7 (SD = 13.02, n=20) at baseline to 23.5 (SD = 15.30, n=17) at 12-weeks. The healthcare worker trial obtained informed consent from 16 participants, 10 commenced therapy and were included in analysis with eight completing therapy. Mean PCL-5 scores decreased from 42.60 (12.23, (n=10) at baseline to 22.00 (19.92, n=8) at 8-weeks.

**Conclusions:** Proof of concept of FIRST was established. PTSD symptom reductions exceeded the PCL-5 minimal clinically important difference. Undertaking a fully powered randomised controlled trial of FIRST therapy is feasible within both healthcare and third sectors.

**Highlights:**

- Post-traumatic stress disorder (PTSD) is more common in military veterans and health workers than the general population
- Therapy can be challenging to commence and complete when it requires a focus on the trauma incident
- FIRST offers a promising, brief, non-trauma focused therapy for the treatment of PTSD

## BACKGROUND

Post-Traumatic Stress Disorder (PTSD) is a mental health condition that may follow experiencing or witnessing traumatic events such as threatened or actual death, serious injury or sexual violence[1]. Numerous occupational groups such as the military, first responders and health and social care workers (HSCWs) are exposed to traumatic events during their working lives, and as such are at increased risk of experiencing PTSD [2, 3]. People who have PTSD experience a significantly decreased quality of life with the condition impacting negatively on day-to-day functioning, physical health and relationships[4–6]. Workers with PTSD are also more likely to perform poorly and/or require sick leave which is an especially important consideration for those in safety critical roles [7–9].

Two populations who frequently experience employment-related PTSD are ex-service personnel of a nation’s armed forces, also known as military veterans [10–12], and HSCWs[4, 13]. We define employment-related PTSD as PTSD resulting from a trauma which has occurred in their workplace or where a work-related event has exacerbated symptoms associated with pre-existing PTSD. The estimated prevalence of PTSD in military veterans is up to 17% in those who served in a combat role during the conflicts in Iraq/Afghanistan[14]; however, we do not know what proportion of veterans with PTSD have work-related PTSD. In some specific contexts military-related PTSD can be as high as 37% in veterans if they continue to live/work in the community where the combat occurred, e.g.Northern Ireland[15]. PTSD in veterans adversely impacts social functioning, increases the likelihood of homelessness and physical health problems, and is frequently associated with mental health comorbidities such as substance misuse and suicidality[1, 5, 16, 17]. It has been found that between 70-80% of treatment-seeking veterans reporting PTSD also have Complex-PTSD (CPTSD). CPTSD results from prolonged or multiple traumas[18]. PTSD and CPTSD are unlikely to resolve spontaneously, and the risk of adverse outcomes increases if untreated [19].

Estimate rates of PTSD in HSCWs lies somewhere between 13% and 45% and this has been exacerbated in those who worked in healthcare through the COVID pandemic[8, 13, 20–22]. In this population, PTSD impacts on workplace performance and sickness absence[7, 9]. Healthcare roles are safety critical; employers also have a legal duty of care to their workforce. Consequently there is a pressing need to find ways to provide evidence-based treatment for HSCWs with PTSD to prevent a steep, and expensive, upward trajectory of healthcare errors, and staffing crises [7, 9]. The reported rates of PTSD in both occupational groups contrast unfavourably with a 5.7% PTSD prevalence in the general English population[23].

The National Institute for Health and Care Excellence (NICE) recommends two evidence-based treatments for PTSD: Trauma Focused Cognitive Behaviour Therapy (TFCBT) and Eye Movement Desensitisation and Reprocessing(EMDR)[24]. However, delivering both therapies requires considerable training, and NICE recommends 8 to 12 sessions as a standard treatment course with more complex cases often requiring considerably more. These are both trauma-focused therapies and treatment completion rates are between 50-85% because toleration for revisiting a trauma therapeutically is challenging[25–27]. As such, scalability of these treatments can pose limitations.

A neurolinguistic programming (NLP)-based, brief, online, cognitive PTSD therapy called Reconsolidation of Traumatic Memories (RTM) has shown promising results in a feasibility randomised controlled trial in 60 UK military veterans undertaken by this team[10]. Subsequently, we modified this NLP therapy to overcome some of the challenges experienced by those presenting within our RTM trial with complex PTSD or with significant dissociation and developed Fast Imagery Reversal Script for Trauma-release^®^ (FIRST^®^). Notably, FIRST is a cognitive therapy with no requirement for the client to recount their trauma story in any detail, and for those with significant dissociation, one of the key steps of the protocol teaches clients how to safely dissociate from their traumatic memories in a controlled manner. The therapy aims to rewrite the emotional elements of the memory and is delivered in 3-4 sessions using a protocolised therapy manual typically over video call. Because of the requirement to adhere closely to a standardised protocol, FIRST can be delivered by a less experienced mental healthcare workforce than the experience required to deliver EMDR or TFCBT offering upscaling advantages in therapist availability without compromising safety and quality. FIRST efficacy is being evaluated in an ongoing efficacy trial with a nested mechanistic experimental study in 215 UK military veterans delivered by the third sector (ISRCTN13985150). This paper presents the results of two proof of concept studies outlining the evidence for the use and evaluation of FIRST in populations with occupationally related PTSD - military veterans and HSCWs.

## RESEARCH DESIGN

Two independent small, single arm, non-controlled proof of concept studies were undertaken in the UK to evaluate acceptability of FIRST and to provide preliminary evidence of an efficacy signal for the impact of FIRST on PTSD symptoms. The two studies are:

**1)** *Veteran PETT* conducted with UK military veterans
**2)** *NHS PETT* conducted with UK HSCW who worked through the COVID-19 pandemic

The feasibility objectives of both studies were to determine whether potentially eligible people would present for FIRST therapy and complete treatment. Efficacy objectives were to determine whether the effect signal found in our previous RTM NLP therapy feasibility trial was maintained with FIRST[10]. Both studies independently included stakeholder involvement: of twelve veterans and fourteen HSCWs, their family members and employers to advise on all aspects of each study and FIRST therapy delivery, respectively.

### FIRST therapy

FIRST therapy is protocolised and delivered in three or four sessions over a minimum of three days and a maximum of four weeks. The protocol is standardised with all populations receiving the same therapy. The number of sessions depends upon how easily the person can develop optimal visualisation skills necessary for the therapy as well as the complexity of PTSD presentation. The two factors determining therapy duration are a) there must be a minimum of one sleep cycle between each therapy session and b) scheduling of therapy sessions for the participant and the therapist. The FIRST protocol therapeutic components and stages are summarised in Table 1.

**Table 1.** The FIRST^®^ Protocol summary and therapeutic suitability.

| Stage | Clinical technique | Theory/evidence for the technique |
| --- | --- | --- |
| Clinical assessment of uncontrolled dissociation mechanisms using the Dissociative Experiences Scale DES-II[28] | The potential participant is assessed to determine their ability to manage their own association and dissociation processes for suitability of the FIRST protocol. | Participants scoring <31 on the DES-II proceed as per therapy protocol.<br>Participants scoring between 32-42* (referred to as High DES-II in following text) proceed with therapist treating them as if the client <u>has</u> a clinically significant level of dissociation and modifies therapeutic protocol. |
| Priming process | Prior to commencing work with a trauma memory, the client is primed to the process by completing a full run through of the key elements of the protocol on a neutral memory experience e.g. making a cup of tea or taking the dog for a walk. | Associational learning provides the opportunity during the priming process to strengthen prefrontal cortex synapses required for memory reconsolidation and alteration in decision making processes[29]. NLP trauma therapy techniques use priming processes before the client engages with the trauma story[30-32].<br>If the client cannot follow the visual format on a neutral activity, FIRST therapy is not suitable for them. |
| Desensitisation process through dissociation | A series of black and white movie repetitions of the trauma event are observed from a twice or doubly dissociated position | FIRST utilises modified variations of black and white movie formats to facilitate controlled dissociation from trauma memory and response[30, 31]. Fear cascade mechanism and the neurobiology of dissociation enable the management of hyperarousal, flashbacks, any derealisation process including dissociation in response to trauma, before the client is required to access an associated memory structure. |
| Undoing of trauma memory through reversed imagery | A fast associated, re-experiencing of the trauma memory in their own body, as a reversed or run backwards imagery of a trauma memory | Individuals with PTSD have a maladaptive response to processing emotions and are stuck in a trauma response that continues to activate a hyperarousal response when certain triggers occur, effectively resulting in the individual continually reliving the trauma experience. By using a fast associated reversed imagery of a trauma memory found in NLP trauma therapies[30, 31], associational learning and adaptive responses are altered and afforded the potential for reconsolidation in the final stage of the processes. |
| Reconsolidation | A new story to the memory is developed which rewrites the emotional elements of the original memory | Rewriting the emotional elements of the memory takes advantage of the hypothesised phenomenon of reconsolidation[33]. Reconsolidation describes the reactivation of long term, otherwise permanent memories, by their evocation in certain contexts. When a memory is reactivated, it labilises, that is, it becomes subject to change. By changing the circumstances surrounding the memory and if the new circumstance provides evidence that a threat of negative emotional stimulus is no longer relevant, the strength of the affective charge may decrease. RTM, used in our previous feasibility trial and sharing characteristics with |
|  |  | FIRST[10], is theorised to use the mechanism of reconsolidation to enable the rewriting of trauma memories. |

\*Veteran PETT eligibility included participants scoring >42

### FIRST therapists

Prospective FIRST therapists working in the NHS or third sector were required to meet criteria of professional standards consistent with their psychological therapy discipline. Therapists were required to hold full or part clinical registration with one of the following recognised professional bodies; British Association for Counselling and Psychotherapy (BACP), the National Counselling and Psychotherapy Society, Accredited Professional Registrant (NCPS Acc.) the British Association of Behavioural and Cognitive Psychotherapy (BABCP), the United Kingdom Council of Psychotherapy (UKCP), the Irish Council for Psychotherapy (ICP). Where partial registration was held, eligibility required completion of core training hours with a UKCP, ICP or BABCP approved training organisation and a minimum of 250 supervised client contact hours. All therapists were required to receive clinical supervision from an accredited supervisor with BACP/UKCP/ICP/BABCP/HCPC. Therapists holding a title of Practitioner Clinical/Counselling Psychologists were additionally required to be registered with the UK Health Care Professions Council (HCPC). All therapists were required to hold professional indemnity insurance with minimum cover of £2.5 million.

### FIRST therapy training

New FIRST therapists completed 10 hours of self-directed learning, undertaken at their own pace, followed by a 2.5-day training via zoom. The training incorporated theoretical teaching, skills-based practice, and structured feedback. During the training each therapist delivered full FIRST protocol with three separate peer ‘clients’. Therapist competency was assessed by the trainer through direct observation of the protocol delivery with a peer client. Therapists had prior experience delivering RTM therapy in our previous trial or had previously been trained in another very similar NLP based trauma treatment protocol[34]. They completed a one-day FIRST-specific training and a single peer-based competency assessment before commencing FIRST delivery in the present study[10]. This training focused on the therapeutic differences between FIRST and the other therapies. Once competency was established, two further fidelity assessments were conducted by a second trainer using a standardised fidelity assessment form to ensure consistency of intervention delivery across all therapists. Therapy sessions were video recorded for the purpose of clinical supervision and protocol assessment fidelity

### Methods: Veteran PETT

This single arm, non-controlled proof of concept (POC) trial evaluated whether FIRST demonstrated efficacy signals comparable to the RTM therapy arm of the prior feasibility trial[10] at 12 weeks with the same number of therapy sessions. Ethical permission was granted by King’s College London Health Facilities (Blue) Research Ethics Committee, ref HR/DP-23/24-41529 on 05/07/2024. Trial registration ISRCTN13985150.

#### Participants and Setting

Participants were UK-based male and female military veterans with a suspected, or confirmed, diagnosis of PTSD and/or CPTSD. Recruitment was conducted via a funded three-month social media campaign previously used successfully[10]. Individuals who screened positive on a study-specific, web-based assessment using the PTSD Checklist for DSM-5 (PCL-5) were invited to provide informed consent[35, 36]. Eligibility was subsequently confirmed during a 90-minute mental health assessment with an Assistant Psychologist trained and supervised in PTSD diagnostics using the Clinician Administered PTSD Scale for DSM-5 (CAPS-5) and the International Trauma Questionnaire (ITQ)[37–39]. The ITQ was used solely to determine the presence of CPTSD. Therapists were recruited from Inspire Wellbeing, a third sector mental healthcare provider in Belfast.

#### Eligibility criteria

Participants were eligible if they 1) were UK based military veterans from the Royal Navy or Royal Marines, British Army or Royal Air Force 2) aged ≥18 years; 3) were currently living or working in the UK; 4) met DSM-5 diagnostic criteria for PTSD as determined by using the CAPS-5 5) experiencing symptoms causing clinically significant distress or impairment in social, occupational or other areas of functioning and 6) consented to therapy sessions being video recorded. Exclusion criteria were 1) currently receiving psychological therapy for PTSD, 2) a comorbid DSM-5 mental disorder of sufficient severity as to interfere with engagement in FIRST therapy, 3) a self-reported suicide attempt within the previous month, 4) currently alcohol dependent determined by an AUDIT cut-off of ≥20[40] or 5) self-reported dependence on prescription or illegal substances or 6) dissociative disorder that moderately or severely impaired function as determined by the Assistant Psychologist during administration of the CAPS-5 assessment.[28].

#### Procedures

Following confirmation of eligibility, demographic information, self-reported PTSD history and baseline data were collected using Qualtrics survey software. Participants were then referred for FIRST commencement within three weeks. Safety assessments were conducted at 6-weeks and follow-up assessments of PTSD and depressive symptoms were completed at 12-weeks (outcomes and measures presented below). In addition, a prespecified battery of mechanisms data proposed for the subsequent efficacy trial was administered at baseline and 12-weeks (mechanistic outcomes not reported here).

#### Sample size

A target sample of 30 participants was selected to match the sample size in the previous feasibility trial[10]. This sample was considered sufficient to estimate the mean change in PTSD symptoms from baseline to 12 weeks, as measured by the PCL-5, with confidence intervals (CI) of comparable width and substantial overlap with the treatment effect signal observed in the RTM arm of the feasibility study.

#### Analysis

Statistical analysis was descriptive and used CI estimates rather than hypothesis testing. Categorical outcomes are reported as numbers and proportions. Continuous outcomes are summarised using descriptive statistics and 95% CIs at each timepoint and for change from baseline. CIs for mean change in outcomes from baseline to 12 weeks were compared with those observed in the previous feasibility study [10]. We also compared the total number of therapy minutes for both therapies to achieve the effect signal.

### Methods: *NHS PETT*

This study employed a pre-RCT preparatory single arm experimental design. Ethical permission was granted by Health Research Authority and Health and Care Research Wales ref: 23/ES/0037 on 04/10/2023. Trial registration ISRCTN52834682.

#### Participants and setting

HSCWs with potential PTSD were referred through their occupational health provider or self-referral to the Staff Counselling and Wellbeing Service (SCaWBs) based at South London and Maudsley NHS Foundation Trust, London. Potentially eligible HSCWs completed a psychological health assessment with SCaWBs staff, after which they were informed about the study and invited to participate. Eligibility was determined by a PCL-5 score <u>≥</u>33[36].

#### Therapist recruitment

Potential FIRST therapists responded to an opportunity to undertake training. The opportunity was promoted to mental and psychological health practitioners supporting SCaWBs with the aim of recruiting and training four staff members.

#### Eligibility criteria

Participants were 1) adults ≥18 years, 2) employed within partnering NHS Trusts, Primary Care or Social Care, between 1st March 2020 and 31st March 2021 covering the three UK waves of the pandemic and 3) diagnosis of PTSD determined by a PCL-5 score ≥33 and confirmed by the NHS Trust Staff Counselling and Wellbeing Service or Post-Incident Pathway (SCaWBs). Ineligible staff were 1) currently receiving psychological therapy for PTSD, 2) unwilling to consent to video-recording of therapy sessions for therapist clinical supervision purposes.

#### Procedures

Baseline data was collected by their allocated therapist using the WriteUpp secure record patient database system following which FIRST therapy commenced within three weeks. Follow up data were collected by SCaWBs at four weeks and eight weeks post baseline. Pseudonymised data were transferred to the research team for data quality monitoring and analysis. Therapists and HSCW participants were invited to participate in a semi-structured online interview with the research team at 8 weeks post baseline. Interviews were audio and visually recorded using Microsoft Teams and underpinned by qualitative realist methodology[41]. Interviews focused on therapists and HSCW participants’ experiences of the delivering or receiving FIRST, the therapy’s mechanistic properties, outcomes and an evaluation of the research procedures.

#### Sample size

A target sample of four psychological wellbeing practitioners and twelve HSCWs was planned to evaluate recruitment, retention and acceptability using progression criteria established in the previous veteran trial[10].

#### Analysis

Frequency and percentages were used to describe participant demographic characteristics, mean and standard deviation for primary and secondary outcomes at individual time-points, and mean differences in PCL-5 from baseline (T1), at final FIRST therapy treatment session (T2), week 4 post-baseline (T3), and week 8 post-baseline (T4) (table 2). The General Anxiety Scale (GAD-7)[42] mean score and standard deviation was calculated for anxiety symptom severity at baseline (T1), week 4 post-baseline (T3), and week 8 post-baseline (T4) and mean differences from baseline (standard deviation) at T3 and T4 as a safety measure. Mean (standard deviation) of secondary outcome scores were calculated at baseline (T1) and week 8 post-baseline (T4) and mean difference (standard deviation) between T1 and T4. Qualitative data were transcribed and pseudonymised by a third party and analysed using reflexive content analysis[43].

**Table 2.** Timing of data collection for person reported outcomes, and measures, of interest.

| Outcomes/<br>follow-up | *Veteran PETT* |  |  | *NHS PETT* |  |  |
| --- | --- | --- | --- | --- | --- | --- |
|  | B/line | 6<br>weeks | 12 weeks | B/line | 4<br>weeks | 8<br>weeks |
| PTSD<br>symptoms<sup>a</sup> | X | X | X | X | X | X |
| Depression<br>symptoms<sup>b</sup> | X | X | X | X | X | X |
| General anxiety<sup>c</sup> |  |  |  | X |  | X |
| Daily life functioning<sup>d</sup> |  |  |  | X |  | X |
| Perceived health status<sup>e</sup> |  |  |  | X |  | X |
| Perceived health status<sup>f</sup> |  |  |  | X |  | X |

Outcomes were assessed by the following <sup>a</sup> PCL-5 <sup>b</sup> PHQ9 <sup>c</sup> GAD7 <sup>b</sup> WSAS <sup>e</sup> EQ-5D-5L <sup>f</sup> EQ-VAS

### Outcomes for Veteran PETT and NHS PETT

#### Feasibility outcomes of interest

In *Veteran PETT*, data were collected on expressions of interest, confirmed participation, therapy commencement and completion with a priori progression criteria. In *NHS PETT* qualitative semi-structured online interviews were conducted with consenting participants and therapists to assess trial and therapy acceptability. For HSCW participants, interviews took place after the final follow-up assessment, whereas therapists were interviewed following completion of therapy delivery to all participants.

#### Clinical outcome

The primary outcome of interest for both studies was PTSD symptom severity assessed by the post-traumatic stress disorder checklist for PTSD symptom severity (PCL-5)[36]. It is a 20-item self-report measure that assesses the 20 DSM-5 symptoms of PTSD typically taking 5-10 minutes to complete. Total scores range from 0-80 with higher scores indicating greater symptom severity. A cut-off score of ≥33 indicates likely PTSD. A 5–10-point change represents reliable change and a ≥10point change represents the minimal clinically significant difference (MCID). The PCL-5 demonstrates high internal consistency in measuring DSM–5 PTSD symptoms[36].

#### Safety outcome

Depressive symptoms were assessed using the Patient Health Questionnaire-9 (PHQ-9), a 9-item self-report measure with each item scored from “0” (not at all) to “3” (nearly every day) [44]. Total scores range from 0-27 and are categorised as follows: 0 to 4 - no significant depressive symptoms; 5 to 9 - mild depressive symptoms; 10 to 14 - moderate; 15 to 19 - moderately severe and 20 to 27 severe depressive symptoms. The PHQ-9 was used to operationalise a safety monitoring protocol, whereby any response other than “0” (not at all) to question 9 triggered a safety contact by the research team.

#### Secondary outcomes of interest in *NHS PETT* only

Secondary outcomes were Work and Social Adjustment Scale (WSAS)[45], the GAD-7[42], and EuroQol five-dimensional five-levels (EQ-5D-5L) and EuroQol-visual analogue scale (EQ-VAS)[46].

## RESULTS

### Veteran PETT

Of 310 expressions of interest, 79 people were initially screened as eligible, 43 attended an eligibility confirmation appointment with the assistant psychologist and twenty participants were confirmed as eligible, consented and enrolled into the study between September 2024 and January 2025. Participants were veterans of the Army (n=14), the Royal Air Force (n=5) and the Royal Marines (n=1).

All participants agreed to take part in FIRST therapy treatment, of whom 17 completed the intervention attending a minimum of three sessions and completed 12-week outcome measures. Participant progression is presented in figure 1. FIRST was delivered by five trained therapists; 13 therapy sessions were assessed for fidelity with at least two sessions included per therapist. All but two sessions for a single therapist met the fidelity threshold; this therapist was removed from the therapist pool during the trial. Participant characteristics are presented in table 4.

**Fig 1.**
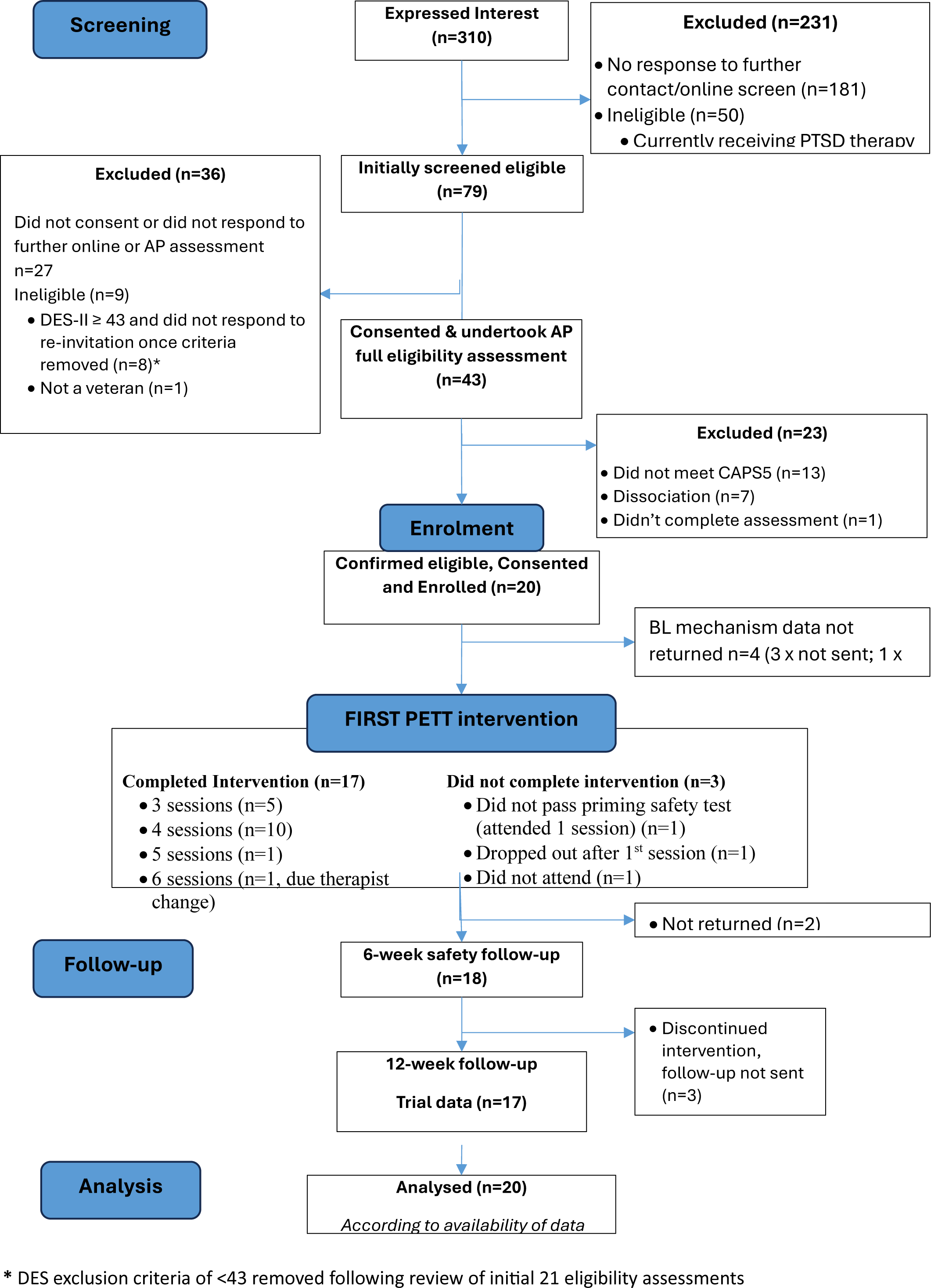
Consort diagram of *Veteran PETT*.

**Table 4.** *VETERAN PETT* Participant characteristics.

| Characteristics | VETERAN PETT<br>(n=20) |
| --- | --- |
| **Sex n (%)** |  |
| Female | 6 (30%) |
| Male | 14 (70%) |
| **Age (years), mean (SD), median** | 44.30 (12.36),<br>42.00 |
| **Ethnicity, n (%)** |  |
| White | 19 (95%) |
| Black African | 1 (5%) |
| **Rank on military exit, n (%)** |  |
| Officer | 8 (40%) |
| Non-commissioned | 12 (60%) |
| **Complex PTSD (ITQ DSO criteria[39]), n (%)** |  |
| Complex PTSD | 16 (80%) |
| Non-complex PTSD | 4 (20%) |
| **Self-reported time since PTSD symptoms onset (years since symptoms started), mean (SD)** | 19.8 (12.84) |
| **Self-reported previous PTSD formal diagnosis, n (%)** | 8 (40%) |
| **Prescribed mental health medication (e.g antidepressants, antipsychotics, sedatives) taken in last 3 months, n (%)** | 8 (40%) |

Mean PCL-5 scores (Table 5) indicated an improvement in PTSD symptoms over time. Scores decreased from a mean of 48.7 (SD = 13.02, n=20) at baseline to 27.2 (SD = 13.61, n=18) at 6 weeks and further to 23.5 (SD = 15.30, n=17) at 12-weeks. These reductions exceeded the 10-point minimal clinically important difference (MCID) for the PCL-5. The mean change from baseline was -21.2 (95% CI –27.9 to -14.4) at 6-weeks and -24.5 (95% CI –32.4 to –16.5) at 12 weeks. At these respective time points, 15 out of 18 participants (83.3%) and 14 out of 17 participants (82.4%) met the threshold for clinically meaningful improvement (10-point difference for the PCL-5), thus meeting the study efficacy signal progression criteria. Figure 2 shows these findings compare favourably to the improvements seen in the previous feasibility trial RTM therapy arm [10].

**Figure 2.**
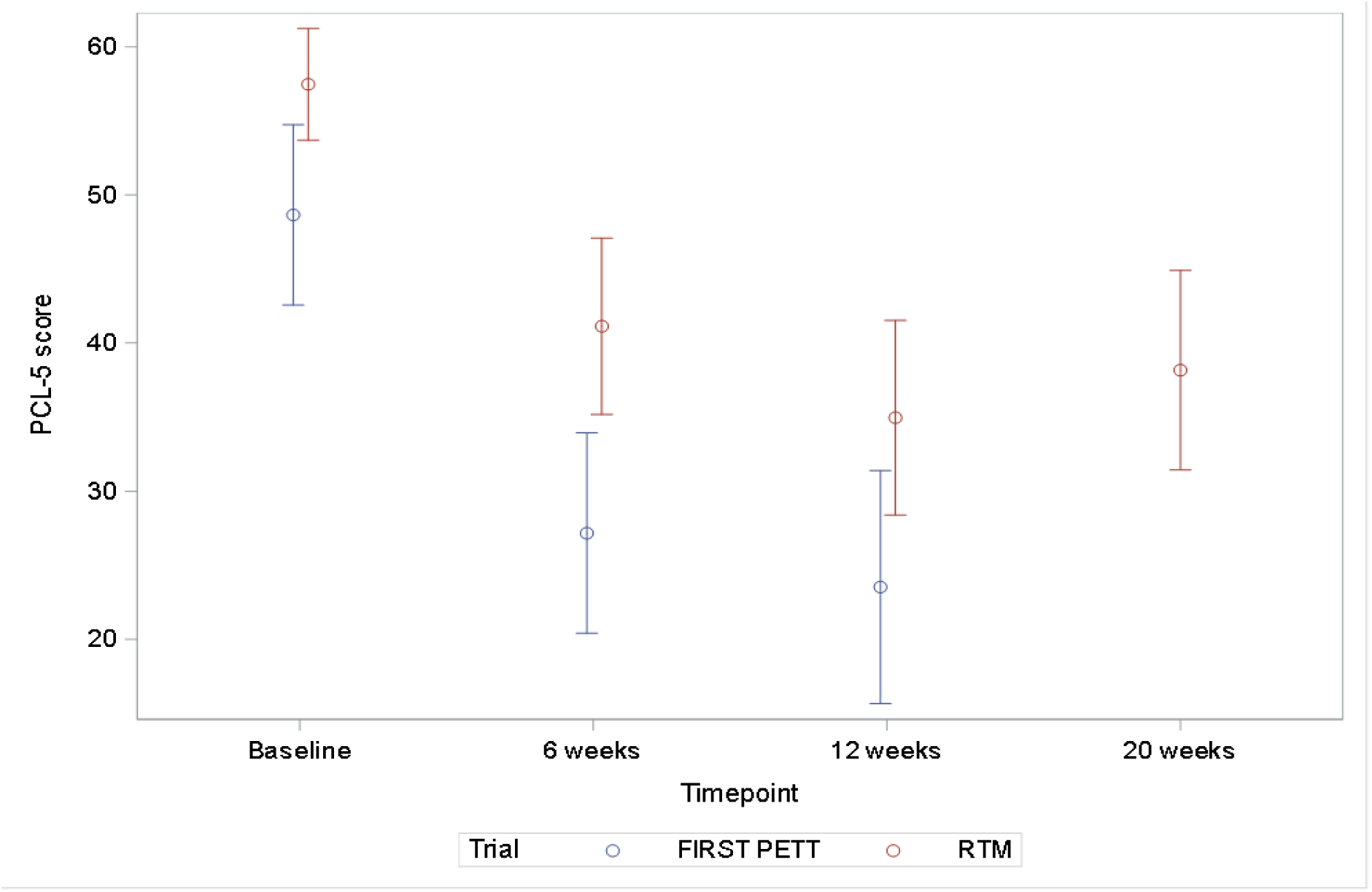
PTSD symptom score **(with unadjusted 95% Confidence intervals)** in the *Veteran PETT* (FIRST PETT) compared to the experimental RTM therapy arm of the prior feasibility study

**Table 5:** *Veteran PETT* FIRST therapy treatment outcome scores at Eligibility/Baseline (T1), Week 6 Post-baseline (T2), and Week 12 Post-baseline (T3) on total sample (n=20)

| Outcome | Baseline (T1) | 6 weeks (T2) | 12 weeks (T3) |
| --- | --- | --- | --- |
| PTSD symptoms (PCL5) Mean (SD), n | 48.7 (13.02), n=20 | 27.2 (13.61), n=18 | 23.5 (15.30), n=17 |
| Change from baseline<br>Mean (SD), n (95% CI) |  | -21.2 (13.61), n=18<br>(-27.9, -14.4) | -24.5 (15.41), n=17<br>(-32.4, -16.5) |
| **MCID ≥10 point**<br>**reduction compared**<br>**to baseline** |  | N=15/18 (83.3%)<br>(60.0%, 95.0%) | N=14/17 (82.4%)<br>(58.2%, 94.6%) |
| Depressive symptoms<br>(PHQ9) |  |  |  |
| Mean (SD), n | 14.6 (5.16), n=19 | 8.9 (5.58), n=18 | 7.9 (7.17), n=17 |
| Change from baseline<br>Mean (SD), n (95% CI) |  | -5.5 (3.71), n=17<br>(-7.4, -3.6) | -7.0 (5.69), n=16<br>(-10.0, -4.0) |

Improvements in depression symptoms were similarly observed on the PHQ-9 with scores decreasing from a mean of 14.6 (SD = 5.16, n=19) at baseline to 8.9 (SD = 5.58, n=18) at 6 weeks and further to 7.9 (SD = 7.17, n=17) at 12-weeks. The safety protocol was triggered for five (15%) participants who responded “Several days” or more to question 9 of the PHQ-9 for whom the research team assessor successfully contacted the participants with no further escalation required (and therefore these instances did not meet criteria for adverse events).

Progression criteria from the POC to the main trial were confirmed with all three pre-specified criteria being met (table 6).

**Table 6.** Feasibility of moving from *Veteran PETT* POC to main trial progression criteria.

| **A priori Progression criteria** | **N (%) participants** | **Rationale** | **Finding** |
| --- | --- | --- | --- |
| Eligible to proceed to receive FIRST therapy | 30 (100%)<br>≥27/30 (90%, ie < 11% STOP)<br><27/30 (90% ie ≥11% STOP) | *In the prior feasibility trial 4/35 (11%) of participants had contraindications to receiving RTM, identified post-randomisation at therapy commencement. We will evaluate our new general mental health eligibility assessment processes (pre-randomisation) to determine participants are FIRST-therapy ready.* | Changes to the screening and recruitment processes from the prior feasibility trial were successful and all recruited participants were found eligible to proceed with therapy.<br>Eligibility progression criteria met. |
| Therapy toleration | ≥25 (83%)<br>15-24 (50-82%)<br><15 (50%) | *PTSD does not resolve itself without treatment so between 15 and 30 participants (50% or over) tolerating therapy offers significant potential treatment gain.* | Post-enrolment, 1/20 (5%) participants were withdrawn from therapy as they did not pass the therapeutic priming process criteria.<br>A total 17/20 (85%) completed therapy, attending a minimum of 3 sessions.<br>Therapy toleration progression criteria met. |
| Efficacy signal – proportion of participants reaching the 10-point MCID reduction at 12 weeks | ≥14 (45%)<br>9 -13 (30-44%)<br><9 (30%) | The feasibility trial found 49% of RTM participants met the 10-point MCID PCL5 reduction at 20 weeks. PTSD does not resolve itself without treatment so with the potential of an efficacious brief therapy an equivalent efficacy signal offers significant potential treatment gain. | 14/17 participants (82%, 95% CI 58%, 95%) reached the 10-point MCID reduction at 12 weeks.<br>Efficacy signal progression criteria met. |

### NHS PETT

Between 22/11/2023 and 25/04/2024, 29 HSCW were assessed for eligibility of which sixteen consented to participate. Four subsequently withdrew prior to therapy commencement, two engaged with one session then withdrew, and ten completed therapy and were included in the final analysis. Two of the 16 scored <32 on the PCL-5 but were symptomatically concerning for the service following assessment and therefore included in the study (figure 3). The mean age of HSCWs was 44 (SD = 12) ranging from 27 to 62 years (Table 7). Eight HSCWs completed four therapy sessions, one had five sessions, and one received three sessions.

**Figure 3:**
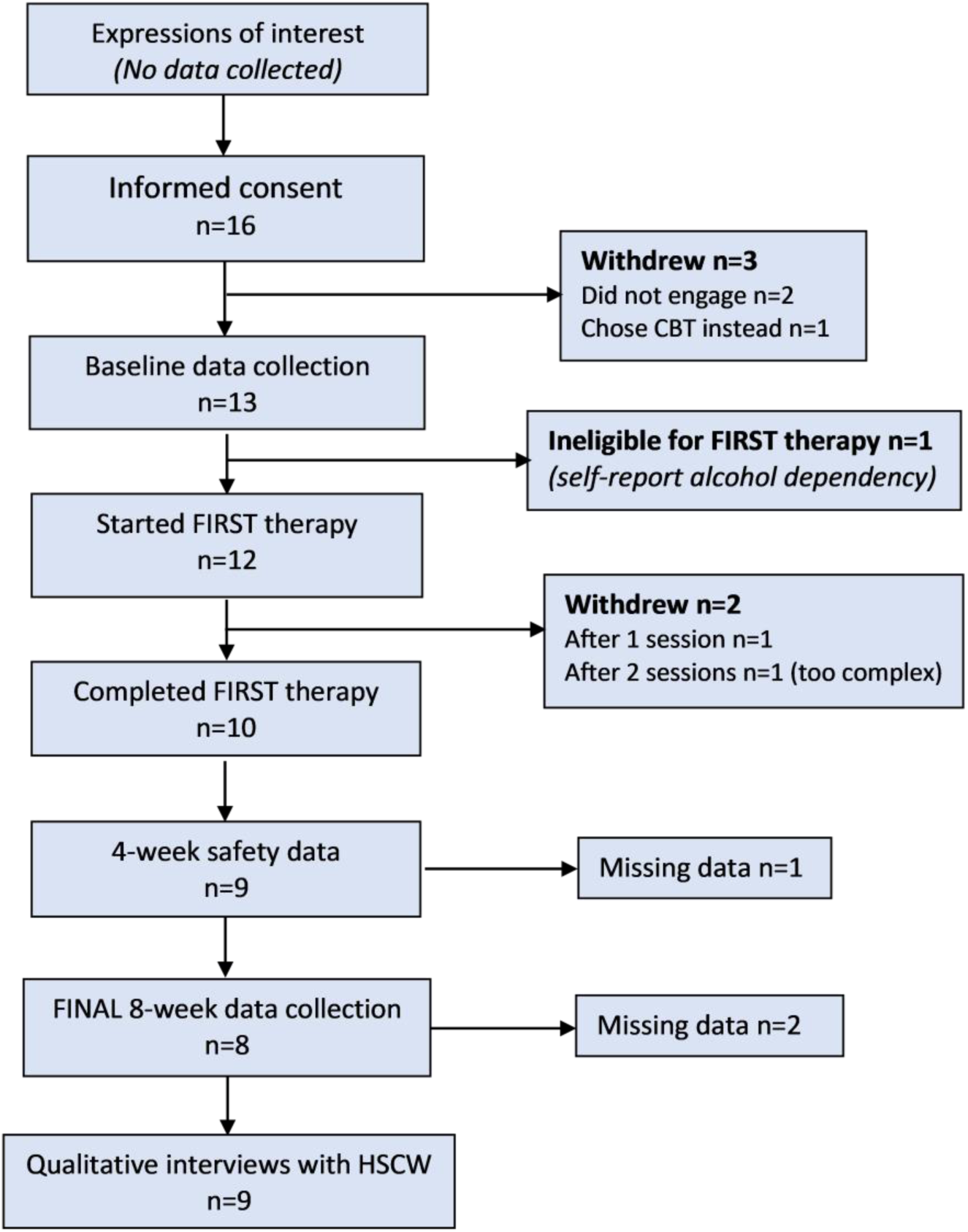
Consort diagram of *NHS PETT*.

**Table 7.**
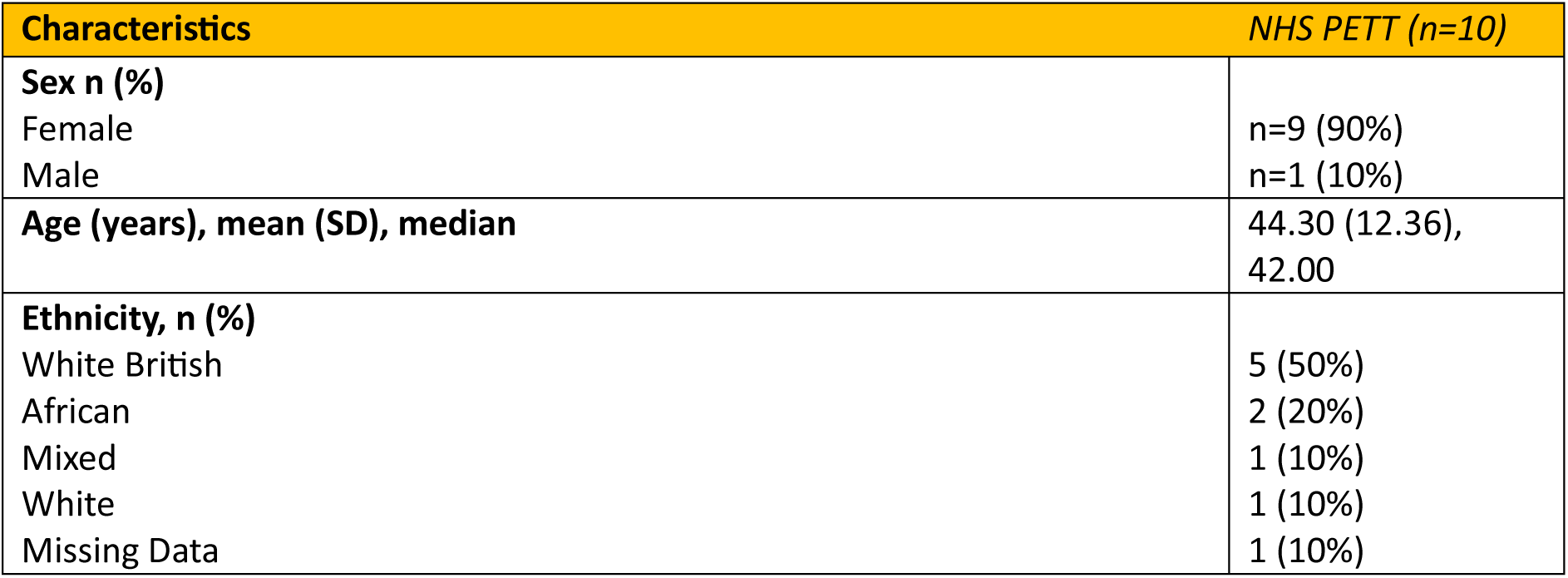
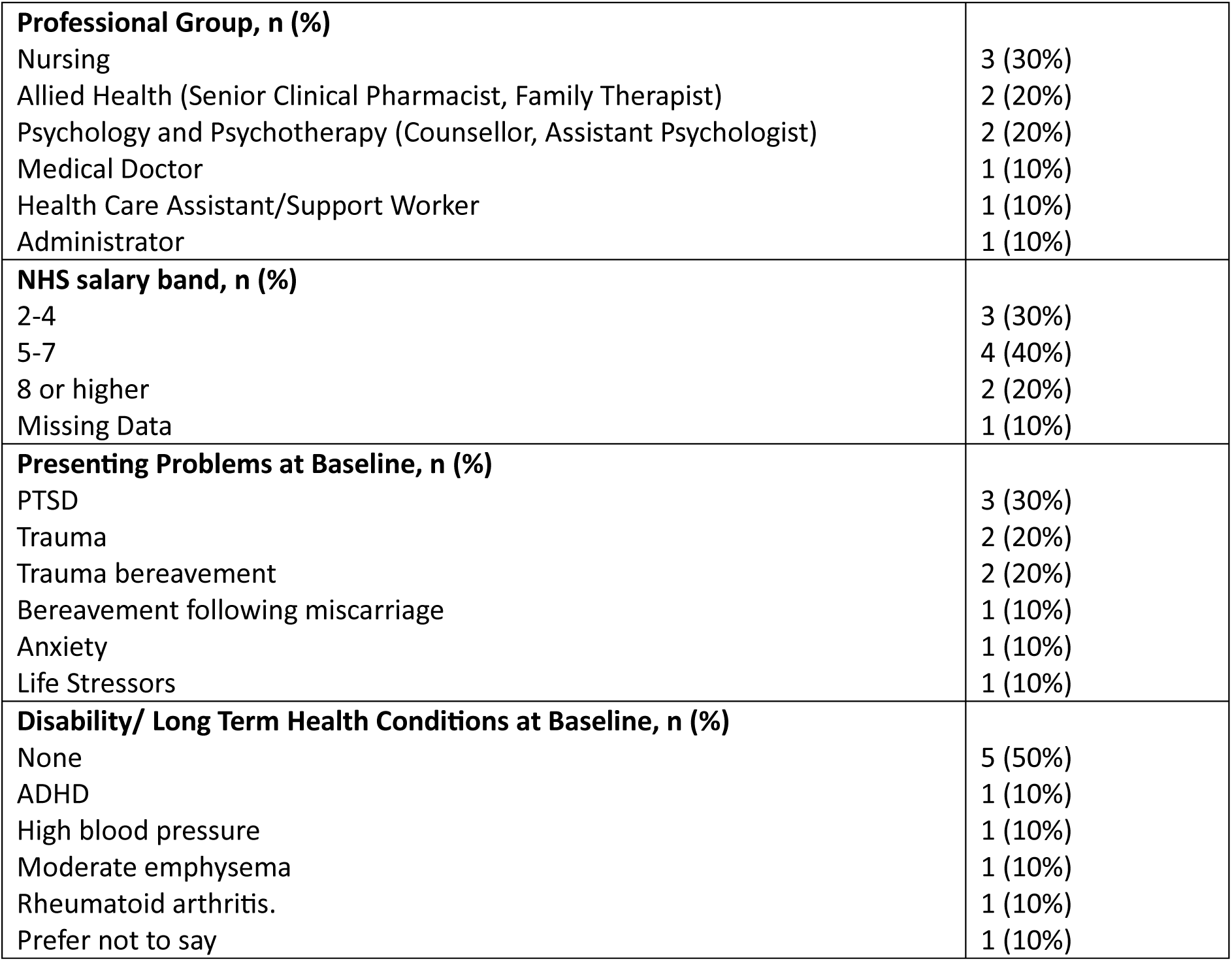
Characteristics of *NHS PETT* participants.

### FIRST^®^ therapy effect signal

Mean PCL-5 scores showed a reduction of greater than the 10-point minimal clinically important difference in PTSD symptoms from baseline to four weeks of 13.38 (SD = 1.09) and baseline to 8 weeks of 20.60 (SD = 7.69). All outcomes improved between baseline and final end point of 8 weeks and are reported in full in table 8. An important contextual improvement to note was the WSAS scores which fell substantially post therapy and exceeded the MCID of 4-5 indicating important increases in social and work functioning[45].

**Table 8:**
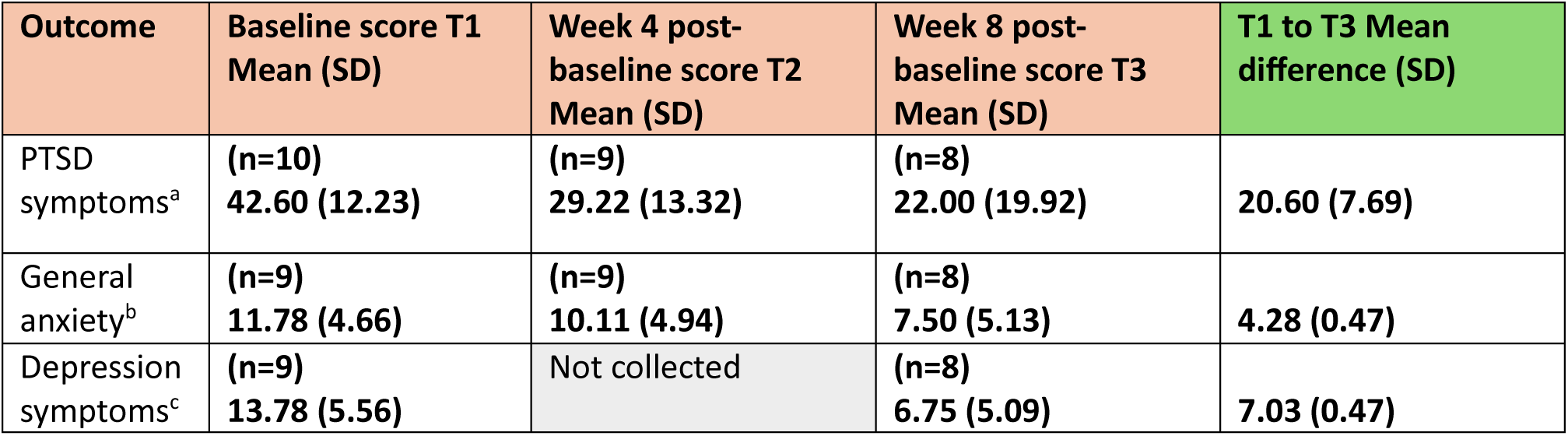
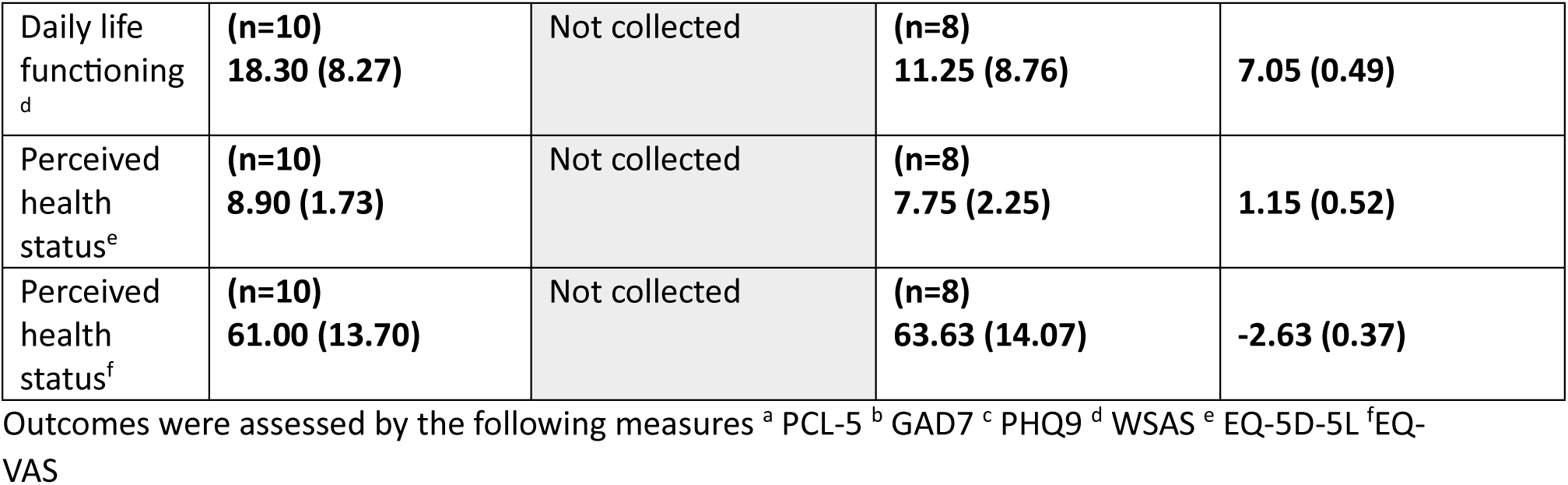
FIRST therapy treatment outcomes at Eligibility/Baseline (T1), Week 4 Post-baseline (T2), and Week 8 Post-baseline (T3) on therapy completers (n=10).

| Outcome | Baseline score T1<br>Mean (SD) | Week 4 post-baseline score T2<br>Mean (SD) | Week 8 post-baseline score T3<br>Mean (SD) | T1 to T3 Mean<br>difference (SD) |
| --- | --- | --- | --- | --- |
| PTSD<br>symptoms<sup>a</sup> | (n=10)<br>42.60 (12.23) | (n=9)<br>29.22 (13.32) | (n=8)<br>22.00 (19.92) | 20.60 (7.69) |
| General<br>anxiety<sup>b</sup> | (n=9)<br>11.78 (4.66) | (n=9)<br>10.11 (4.94) | (n=8)<br>7.50 (5.13) | 4.28 (0.47) |
| Depression<br>symptoms<sup>c</sup> | (n=9)<br>13.78 (5.56) | Not collected | (n=8)<br>6.75 (5.09) | 7.03 (0.47) |
| Daily life functioning<sup>d</sup> | (n=10)<br>18.30 (8.27) | Not collected | (n=8)<br>11.25 (8.76) | 7.05 (0.49) |
| Perceived health status<sup>e</sup> | (n=10)<br>8.90 (1.73) | Not collected | (n=8)<br>7.75 (2.25) | 1.15 (0.52) |
| Perceived health status<sup>f</sup> | (n=10)<br>61.00 (13.70) | Not collected | (n=8)<br>63.63 (14.07) | -2.63 (0.37) |

Outcomes were assessed by the following measures <sup>a</sup> PCL-5 <sup>b</sup> GAD7 <sup>c</sup> PHQ9 <sup>d</sup> WSAS <sup>e</sup> EQ-5D-5L <sup>f</sup> EQ-VAS

Participant safety was successfully monitored at each therapy treatment. There were no adverse events or serious adverse events reported in this study. None of the HSCW participants met our safety criteria relating to changes in PTSD symptoms from the PCL-5 mean scores between sessions or from baseline to session.

### Feasibility of FIRST to therapists

Seven NHS therapists requested training as FIRST therapists. Five met the criteria for training and commenced, with four completing training. One therapist was unable to commit to the training time requirements. Participating therapists were a social worker, an occupational therapist and two cognitive behaviour therapists. Qualitative interviews with therapists revealed that training was acceptable and prepared them for delivering FIRST within the NHS (table 9). They found video-call/online training to be flexible and acceptable, and group learning with peers was advantageous. The three-day FIRST training was reported as intense, insightful and informative. Negative experiences related to supervisory feedback and information density. The protocolised nature of FIRST left therapists uncomfortable with not having access to client general mental health and trauma history. Therapists familiar with high intensity protocolised therapies reported developing FIRST competency quickly. Therapists skilled in holistic client work reported struggling to focus entirely on the PTSD.

**Table 9:** Acceptability of FIRST Training: *NHS PETT* study therapists.

| FIRST training | Acceptability description | Quotation |
| --- | --- | --- |
| Overall training feedback | The training was acceptable. Therapists found it insightful and positively challenging, though the workload alongside regular work commitments felt demanding. Online delivery facilitated flexibility, but intensity and time commitment were overwhelming at times. | "I definitely understood the time commitment... all of that was very clear... the reality of training... on top of your day job meant...[you] don't have time to do anything, so like adjusting your diary... was a lot" (NHSPETT020).<br>"The training was probably a little bit more intense than I had bargained for, it was a learning curve" (NHSPETT017). |
| Peer learning | Peer training was acceptable and valued for shared context and mutual understanding. Peer supervision was initially helpful, though the relevance of the peer supervision reduced as therapists progressed at different speeds. | "I think it was good training with your peers really because also we all come from the same sort of working environment, so when we were using practice examples you could understand where they were coming from or that felt familiar" (NHSPETT021).<br>"I quite liked the group supervision...but it didn't stay as a group supervision for very long" (NHSPETT017). |
| 1-2-1 supervision | Individual supervision was acceptable and viewed as supportive. It helped therapists consolidate learning and reflect on practice and delivery | "I thought the supervision was good... very helpful" (NHSPETT017).<br>"It allowed time to reflect" (NHSPETT020). |
| Practice clients | Using practice clients was acceptable and useful for developing skills. However, some noted that the practice clients were overly compliant and not fully representative of real-world patient presentations. | "[Practice clients] and recording them and getting them critiqued... really helpful... because there's so many little things that you don't realise you're doing at the beginning" (NHSPETT019).<br>"The two practice [clients], which was really helpful... but they're very compliant... You learn a lot about your own skills... like building confidence (NHSPETT018). |
| Protocolisation | Acceptability of the FIRST protocol was mixed. Some found its structure reassuring and containing, particularly those accustomed to protocolised work. Other therapists experienced the protocol as inflexible, limiting rapport-building and leaving them feeling deskilled. | "It's fairly predictable what will happen" (NHSPETT019).<br>"It was incredibly containing" (NHSPETT021).<br>"I felt actually some sort of reassurance" (NHSPETT020).<br>"There's not much space to build rapport, because you basically have to follow the script" (NHSPETT018).<br>"I just felt it quite deskilling...so I just had to read from a script" (NHSPETT018). |

### Feasibility of FIRST to HSCW with PTSD

FIRST therapy was acceptable to most participating HSCW with PTSD (table 10). Five experienced initial challenges during the first two sessions and said time was needed post session to process before returning to work. Two HSCWs found FIRST unacceptable because they wanted a talking therapy. Despite some facing technological challenges, therapy via video call was acceptable. The disadvantage for two was the reduced human connection. Most HSCWs found the practice visualisations acceptable in preparation for therapy however these visualisations were not without challenges and HSCWs continued to find these challenging into the main visualisation phase, as they struggled to stay on task, deploy some of the necessary cognitive strategies and disliked visualising themselves experiencing distress. Overall, FIRST therapy was described as mentally draining, exhausting and depleting. Whilst there were mixed responses to use of a strict therapy protocol and a lack of talking, over time HSCWs became accustomed to the techniques and reported positive relationships with their therapist, describing them as empathetic, engaging, accommodating and calming. Initial reservations about therapy brevity were overcome as symptoms diminished.

**Table 10:** Acceptability of the FIRST therapy *NHS PETT* study clients.

| FIRST delivery | Acceptability | Quotation |
| --- | --- | --- |
| Overall delivery | First was generally acceptable to clients, though the initial two sessions were described as draining and fatiguing. Fatigue eased as therapy progressed. While clients initially question whether a four-session intervention would be sufficient prior to treatment, following treatment most found the brevity acceptable due to noticeable progress and symptom reduction. | "...The first two sessions were hard... Yeah, quite exhausted and drained...less so in the most recent two. But after the first two, yeah, I was super, super tired" (NHSPETT013).<br>"I was worried that...I would be left after the 4 weeks feeling quite distressed...like I hadn't covered everything and still feeling like I needed support" (NHSPETT006)<br>"I made such good progress week on week that actually really helped... It's kind of quite intense but it hits the spot" (NHSPETT008). |
| Online delivery | Online delivery was acceptable for most clients, particularly as it supported attendance around workload and their personal commitments. However, some clients experienced technical difficulties and struggled to build rapport with their therapist online. | "I mean, it worked for me... doing it remotely... that facilitated my ability to attend to every session" (NHSPETT008).<br>"...Being on Teams it's a bit difficult to establish that relationship" (NHSPETT007).<br>"We both at times had real issues with internet connection and network issues" (NHSPETT008). |
| Protocolisation | The structured protocol had mixed acceptability. Some clients found the predictability and clear structure reassuring, while others experienced it as rigid, robotic and less conducive to emotional sharing. | "I work quite well with structure and knowing what's coming and yeah, so I think for me that was helpful" (NHSPETT006).<br>"It was very protocol based - I think that was a bit difficult for me... less of the protocol and more of the human...because you are talking about your emotions...I found that disorientating..." (NHSPETT007). |
| Practice visualisations | The practice visualisations were acceptable and helped clients become familiar with the techniques used in the main therapy session. | "I thought it would be very difficult but actually after a few practices - I don't know if this was a common experience, but...I felt like it was quite quick to get" (NHSPETT006). |
| Visualisations | Visualisation tasks were challenging for many clients, particularly the speed and technical demands of reversing and replaying scenes in the mind. Many thought this was too cognitively demanding. Other found the techniques helpful, grounding and comforting, with some continuing to use the techniques outside the therapy session. | "I am quite a visual person but I really struggled to do that" (NHSPETT001).<br>"Difficult, because two seconds is nothing at all. But I found that difficult more logistically than emotionally weirdly, because it was just so hard to get it in the two seconds...it was tiring. It wasn't pleasant" (NHSPETT013).<br>"I went back to the same booth... I found that safe almost" (NHSPETT006). |
| Relationship with therapist | Most clients reported a positive therapeutic relationship, describing their therapist as empathetic, calm and supportive. A minority experienced difficulty in forming rapport, particularly online and with a rigid protocol. | "I felt an initial kind of rapport with her, I think she was really sensitive and empathetic" (NHSPETT008).<br>"She was very empowering and encouraging" (NHSPETT001).<br>"I didn't feel that necessarily I was making any contact with her as a human being." (NHSPETT003). |

## DISCUSSION

We propose that FIRST therapy proof of concept is confirmed in both populations for detecting an efficacy signal, determining acceptability of trial procedures and training a range of mental healthcare staff.

An efficacy signal was observed, with a mean reduction of PTSD symptoms on the PCL-5 of 25-points in military veterans 12 weeks from baseline and 21-points in healthcare workers eight weeks from baseline. These reductions exceed the 10-point MCID and provide preliminary evidence of efficacy to be evaluated in further controlled trials. These results compared well with our previous evaluation of NLP-based therapy, RTM, of 60 veterans which showed an 18-point reduction at 20-weeks compared to an 8-point reduction in the TFCBT group[10]. The fall in PCL5 scores also compares favourably with previous trials of TFCBT and EMDR in other populations [1]. A service evaluation of NHS Talking therapies reports on 1580 people being treated over 11 years found reductions of 18-points associated with both TFCBT and EMDR [25]. These results were obtained in therapy of 3-4 FIRST sessions compared to the much longer treatment duration of TFCBT and EMDR [1]. No adverse events were reported in either study.

Both populations in this study routinely face challenges engaging with trauma-focused therapy, which can contribute to dropout [27]. Therapy completion was 17/20 (85%) in *Veteran PETT* and in 10/16 (63%) *NHS PETT,* within the typical ranges 50-85% reported for trauma focused interventions [25–27]. These findings highlight the ongoing need to ensure therapy feels safe and trustworthy across PTSD populations. The *Veteran PETT* recruitment target of 30 participants was not met due to higher than anticipated exclusions based on their DES score during the AP assessments. The eligibility criteria around dissociation and the DES score illustrated in figure 1 has since been revised in the ongoing main trial.

We have demonstrated that statutory and third sector mental health staff, meeting minimal eligibility criteria, can be trained to deliver FIRST therapy, via video call, to small groups of HSCWs and ex-service personnel with PTSD. Qualitative findings highlight challenges in delivering protocolised therapy with limited client history. While delivery was feasible, having prior client history was preferable to better understand and support participants. Improvements in client symptomatology may make FIRST increasingly acceptable over time. Expectation preparation may be required in the future so potential FIRST therapists have a better understanding of this context before embarking upon training. Informal stakeholder discussions with third sector therapists mirrored these themes.

The two populations differed in sex and baseline PTSD severity. Most HSCWs were female, while most military veterans were male. Despite likely differences in primary trauma, FIRST demonstrated a strong efficacy signal across both sexes. Nine of the ten HSCWs were female versus 74% of the NHS workforce being female [47]. In *Veteran PETT*, 80% (16/20) met criteria for complex PTSD, this was not assessed in the HSCWs. A PCL-5 cut-off of 51 has been proposed to indicate complex PTSD, suggesting a higher prevalence in veterans [48]. *Veteran PETT* participants represented multiple military services, predominantly army veterans, with a wide age range. *NHS PETT* participants were diverse in age, ethnicity, disability, pay bands and clinical specialities.

### Study strengths and limitations

These studies were conducted in contrasting settings, with therapists working inside and outside the NHS demonstrating proof of concept for upscaling FIRST therapy for PTSD via multiple delivery routes. Both clinical providers operated in pragmatic, high-demand settings with rapid mental healthcare assessments and treatment waiting lists. In both organisations service changes were required to implement FIRST and organisational flexibility enabled delivery of FIRST training and therapy within the trial timelines. Whilst efforts have been taken to present the methods and results of the two separate studies in ways which optimise comparability, key methodological differences remain which could be more robustly addressed in a single trial encompassing both populations. The PCL5 has demonstrated validity and reliability across trauma populations supporting its use as a robust comparative measure [49].

Both studies were underpowered, uncontrolled, and subject to participant eligibility constraints. Regression to the mean may partly explain observed reductions in PTSD symptomatology. Ongoing full trials are required to determine efficacy and to test mechanistic hypotheses regarding how FIRST exerts its effect.

## CONCLUSIONS

Our findings suggest that a fully powered randomised controlled trial of FIRST^®^ therapy is feasible within both the NHS and the third sector settings. POC was demonstrated in 30 participants across two distinct PTSD populations. Future research should evaluate FIRST in broader populations including first responders, police, border control personnel, social workers. Conducting a large pragmatic trial comparing the cost-effectiveness of FIRST with current gold standard therapies could bring significant evidential benefits to the field and ultimately to both employees and their employers.

## Data Availability

The data that support the findings of this study are available from the corresponding author upon reasonable request

## Acknowledgements

We would like to thank the public contributors from the veteran and the NHS communities who supported these studies, the therapists who embraced the opportunity to deliver an experimental therapy and the participants, without whose commitment to their own mental health and that of their wider communities, this research would not be possible.

